# Biophysical Characterization of A Novel *SCN5A* Mutation Associated with an Atypical Phenotype of Atrial and Ventricular Arrhythmias and Sudden Death

**DOI:** 10.1101/2020.09.04.20186171

**Authors:** Mohammad-Reza Ghovanloo, Joseph Atallah, Carolina A. Escudero, Peter C. Ruben

## Abstract

**Background:** Sudden cardiac death (SCD) is an unexpected death that occurs within an hour of the onset of symptoms. Hereditary primary electrical disorders account for up to 1/3 of all SCD cases in younger individuals and include conditions such as catecholaminergic polymorphic ventricular tachycardia (CPVT). These disorders are caused by mutations in the genes encoding cardiac ion channels, hence they are known as cardiac channelopathies. We identified a novel variant, T1857I, in the C-terminus of Nav1.5 (*SCN5A*) linked to a family with a CPVT-like phenotype characterized by atrial tachy-arrhythmias and polymorphic ventricular ectopy occurring at rest and with adrenergic stimulation, and a strong family history of SCD.

**Objective:** Our goal was to functionally characterize the novel Nav1.5 variant and determine a possible link between channel gating and clinical phenotype.

**Methods:** We first used electrocardiogram recordings to visualize the patient cardiac electrical properties. Then, we performed voltage-clamp of transiently transfected CHO cells. Lastly, we used the ventricular/atrial models to visualize gating defects on cardiac excitability.

**Results:** Voltage-dependences of both activation and inactivation were right-shifted, the overlap between activation and inactivation predicted increased window currents, the recovery from fast inactivation was slowed, there was no significant difference in late currents, and there was no difference in use-dependent inactivation. The O’Hara-Rudy model suggests ventricular afterdepolarizations and atrial Grandi-based model suggests a slight prolongation of atrial action potential duration.

**Conclusion:** We conclude that T1857I likely causes a net gain-of-function in Nav1.5 gating, which may in turn lead to ventricular afterdepolarization, predisposing carriers to tachy-arrhythmias.

## Introduction

Sudden cardiac death (SCD) is an unexpected death that occurs within an hour of the onset of symptoms^1,2^. SCD is a devastating public health problem with major social implications and is responsible for thousands of deaths annually worldwide^1^. Hereditary primary electrical disorders account for up to a third of all SCD cases in younger individuals^3^ and include: long QT syndrome (LQTS), Brugada syndrome (BrS), short QT syndrome (SQT), and catecholaminergic polymorphic ventricular tachycardia (CPVT)^4–6^. These disorders are caused by mutations in the genes that encode cardiac ion channels, hence they are known as cardiac channelopathies.

CPVT is a malignant inheritable cardiac channelopathy that is triggered by stress or exertion and is characterized by atrial tachycardia, polymorphic ventricular arrhythmias and SCD. Most mutations in patients with CPVT have been reported in the gene that encodes the cardiac ryanodine receptor (*RyR2*). In addition to *RyR2*, few cases of CPVT have been caused by mutations in the genes that encode calsequestrin (*CASQ2*) and Kir2.1 (*KCNJ2*)^4,5^.

The sodium current passing through voltage-gated sodium channels (Nav) initiates action potentials in excitable cells. Nav channels are hetero-multimeric proteins composed of large ion conducting α-subunits and smaller auxiliary β-subunits^7–12^. The *SCN5A* gene encodes Nav1.5, which is the predominantly expressed Nav in the myocardium^13^. *SCN5A* mutations have been associated with LQTS, BrS, cardiac conduction defects, sick sinus syndrome, and dilated cardiomyopathy^14^.

This study focuses on a novel mutation, T1857I, in the C-terminus of Nav1.5 that is linked to a family with an inherited arrhythmogenic phenotype characterized by atrial tachy-arrhythmias and polymorphic ventricular arrhythmia occurring at rest and with adrenergic stimulation, and a strong family history of SCD, akin to but not typical of CPVT. We show full biophysical characterization of this mutation using whole-cell voltage-clamp.

## Materials and Methods

### Cell Culture

Chinese Hamster Ovary (CHO) cells were transiently co-transfected with cDNA encoding eGFP and the β1-subunit and either WT or variant Nav1.5 α-subunit. Transfection was done according to the PolyFect transfection protocol. After each set of transfections, a minimum of 8-hour incubation was allowed before plating on sterile coverslips.

### Electrophysiology

Whole-cell patch-clamp recordings were performed in an extracellular solution containing (in mM): 140 NaCl, 4 KCl, 2 CaCl_2_, 1 MgCl_2_, 10 HEPES (pH 7.4). Solutions were adjusted to pH7.4 with CsOH. Pipettes were filled with intracellular solution, containing (in mM): 120 CsF, 20 CsCl, 10 NaCl, 10 HEPES. All recordings were made using an EPC-9 patch-clamp amplifier (HEKA Elektronik, Lambrecht, Germany) digitized at 20 kHz via an ITC-16 interface (Instrutech, Great Neck, NY, USA). Voltage-clamping and data acquisition were controlled using PatchMaster software (HEKA Elektronik, Lambrecht, Germany) running on an Apple iMac. Current was low-pass-filtered at 10 kHz. Leak subtraction was performed automatically by software using a P/4 procedure following the test pulse. Gigaohm seals were allowed to stabilize in the on-cell configuration for 1 min prior to establishing the whole-cell configuration. Series resistance was less than 5 MΩ for all recordings. Series resistance compensation up to 80% was used when necessary. All data were acquired at least 1 min after attaining the whole-cell configuration. Before each protocol, the membrane potential was hyperpolarized to −130 mV to ensure complete removal of both fast inactivation and slow inactivation. All experiments were conducted at 22 °C.

### Activation Protocols

To determine the voltage-dependence of activation, we measured the peak current amplitude at test pulse potentials ranging from −100 mV to +80 mV in increments of +10 mV for 20 ms. Channel conductance (G) was calculated from peak I_Na_:

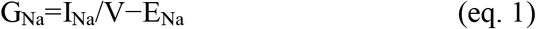

where G_Na_ is conductance, I_Na_ is peak sodium current in response to the command potential V, and E_Na_ is the Nernst equilibrium potential. Calculated values for conductance were fit with the Boltzmann equation:

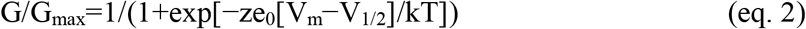

where G/G_max_ is normalized conductance amplitude, V_m_ is the command potential, z is the apparent valence, e_0_ is the elementary charge, V_1/2_ is the midpoint voltage, k is the Boltzmann constant, and T is temperature in K.

### Steady-State Fast Inactivation Protocols

The voltage-dependence of fast inactivation was measured by preconditioning the channels to a hyperpolarizing potential of −130 mV and then eliciting pre-pulse potentials that ranged from −170 to +10 mV in increments of 10 mV for 500 ms, followed by a 10 ms test pulse during which the voltage was stepped to 0 mV. Normalized current amplitudes from the test pulse were fit as a function of voltage using the Boltzmann equation:

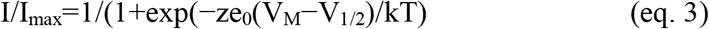

where *I*_max_ is the maximum test pulse current amplitude.

### Recovery from Fast Inactivation Protocols

Channels were fast-inactivated during a 20 ms or 200 ms depolarizing step to 0 mV, and recovery was measured during a 19 ms test pulse to 0 mV following a −90 mV recovery pulse for durations between 0 and 1.024 s. Time constants of fast inactivation recovery showed two components and were fit using a double exponential equation:

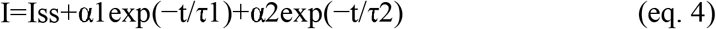

where *I* is current amplitude, *I*_ss_ is the plateau amplitude, α_1_ and α_2_ are the amplitudes at time 0 for time constants τ_1_ and τ_2_, and *t* is time.

### Use-Dependent Inactivation - 3 Hz

Channels accumulated into a use-dependent inactivated state during either a series of depolarizing pulses to 0 mV. Normalized current amplitude as a function of time was fitted with a double exponential.

### Action Potential Modeling

Ventricular action potentials were simulated using a modified version of the O’Hara– Rudy model programmed in Matlab^15,16^. The code that was used to produce the model is available online from the Rudy Lab website (http://rudylab.wustl.edu/research/cell/code/Allcodes.html). The modified gating INa parameters were in accordance with the most relevant biophysical data obtained from whole cell patch clamp experiments in this study for various conditions. Atrial action potentials were simulated using a modified version of the Grandi model coded in Matlab^17,18^. The original code was acquired from UC Davis (https://somapp.ucdmc.ucdavis.edu/Pharmacology/bers/).

### Analysis

Analysis and graphing were done using FitMaster software (HEKA Elektronik) and Igor Pro (Wavemetrics, Lake Oswego, OR, USA). All data acquisition and analysis programs were run on an Apple iMac (Apple Computer). Statistical analysis was performed in JMP version 13.

### Statistics

Continuous variables are presented as means ± standard deviation and were normally distributed. T-test was used to compare the mean responses [activation, current density, steady-state fast inactivation, late currents, use-dependent inactivation, and fast inactivation recovery] between channel variants (two levels: WT and T1857I). A level of significance α = 0.05 was used in all overall tests, and effects with p-values less than 0.05 were considered statistically significant.

## Results

### Phenotypic characterization

The T1857I mutation was identified in siblings. There was no history of arrhythmic syncope or seizure. The family history was relevant for SCD in one parent and one grandparent, both occurring at rest. The remaining maternal family history is relevant for multiple individuals with early onset atrial fibrillation and SCD. Investigations included serial ECGs, signal average ECGs, high-lead ECGs, exercise stress tests, 24-hour Holter monitoring, echocardiograms, and cardiac magnetic resonance imaging. These investigations demonstrated a pattern of progressively increasing burden of atrial and ventricular ectopy and non-sustained tachycardia of post-pubertal onset. The arrhythmias worsened with exercise but were also ambient at baseline. Criteria were not met for any cardiac channelopathy, specifically a normal QTc at baseline and in recovery on exercise stress test, no evidence of Brugada like changes, no pathologic early repolarization, as well as no evidence of cardiomyopathy. The clinical characteristics were suggestive of an atypical CPVT-like phenotype. A broad arrhythmia genetic screening panel was performed, identifying the novel T1857I mutation in both siblings.

During 5 years of follow-up, both siblings developed a very similar phenotype with frequent catecholamine sensitive atrial and polymorphic ventricular ectopy with non-sustained atrial tachycardia up to 200 bpm along with bidirectional PVCs and polymorphic triplets. The arrhythmias are only partially responsive to Nadolol and were thought to be minimally responsive to Flecainide. Sample ECG is shown in **Figure 1** showing sinus rhythm with frequent polymorphic premature wide complex beats, depolarization abnormality with low T-wave amplitude and inverted T-waves in the right precordial leads (V1 and V2), and otherwise normal intervals.

**Figure 1.**
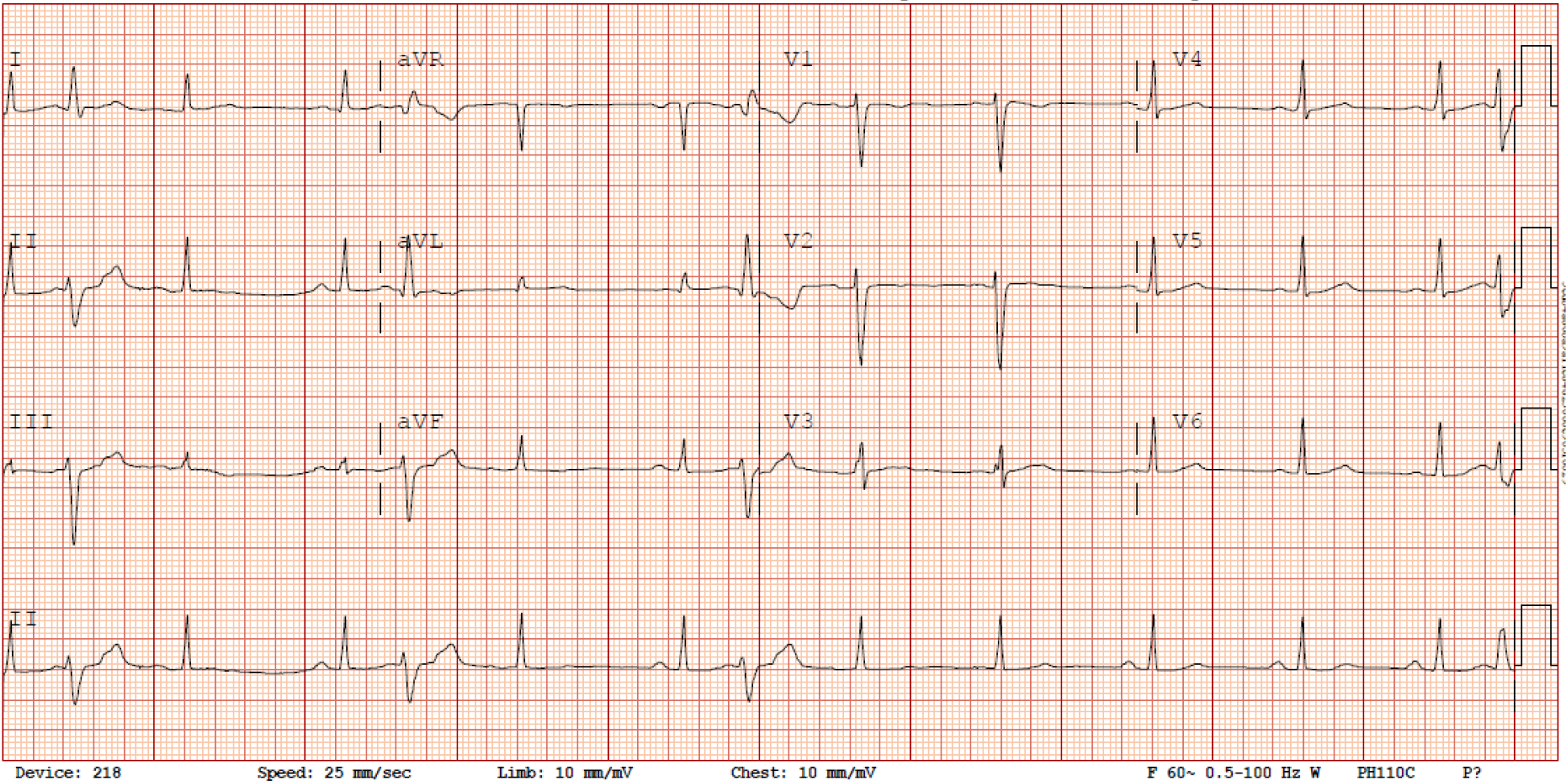
– Standard 12-lead ECG obtained from the patient with the novel *SCN5A* mutation. Sinus rhythm with frequent polymorphic premature wide complex beats, low T-wave amplitude and inverted T-waves in the right precordial leads (V1 & V2), normal axis and QRS duration and QTc = 396.

### T1857I Depolarizes Activation, Increases Apparent Valence

We conducted whole-cell patch-clamp experiments to investigate whether the T1857I variant in the C-terminus alters the biophysical properties of Nav1.5. We examined the variant effect on channel activation in WT and T1857I by measuring peak channel conductance at membrane potentials between −100 and +80 mV (**Figure 2A-B**). We found that the mutation causes a significant shift on the midpoint (V1/2) of the conductance curve (p=0.0069) in the depolarized direction. T1857 also causes a significant increase in the apparent valence (z) of activation (p=0.0317) (**Table S1**). This suggests that this C-terminal mutation has a destabilizing effect on the voltage-dependence of activation and also affects magnitude of charge movement during activation.

**Figure 2.**
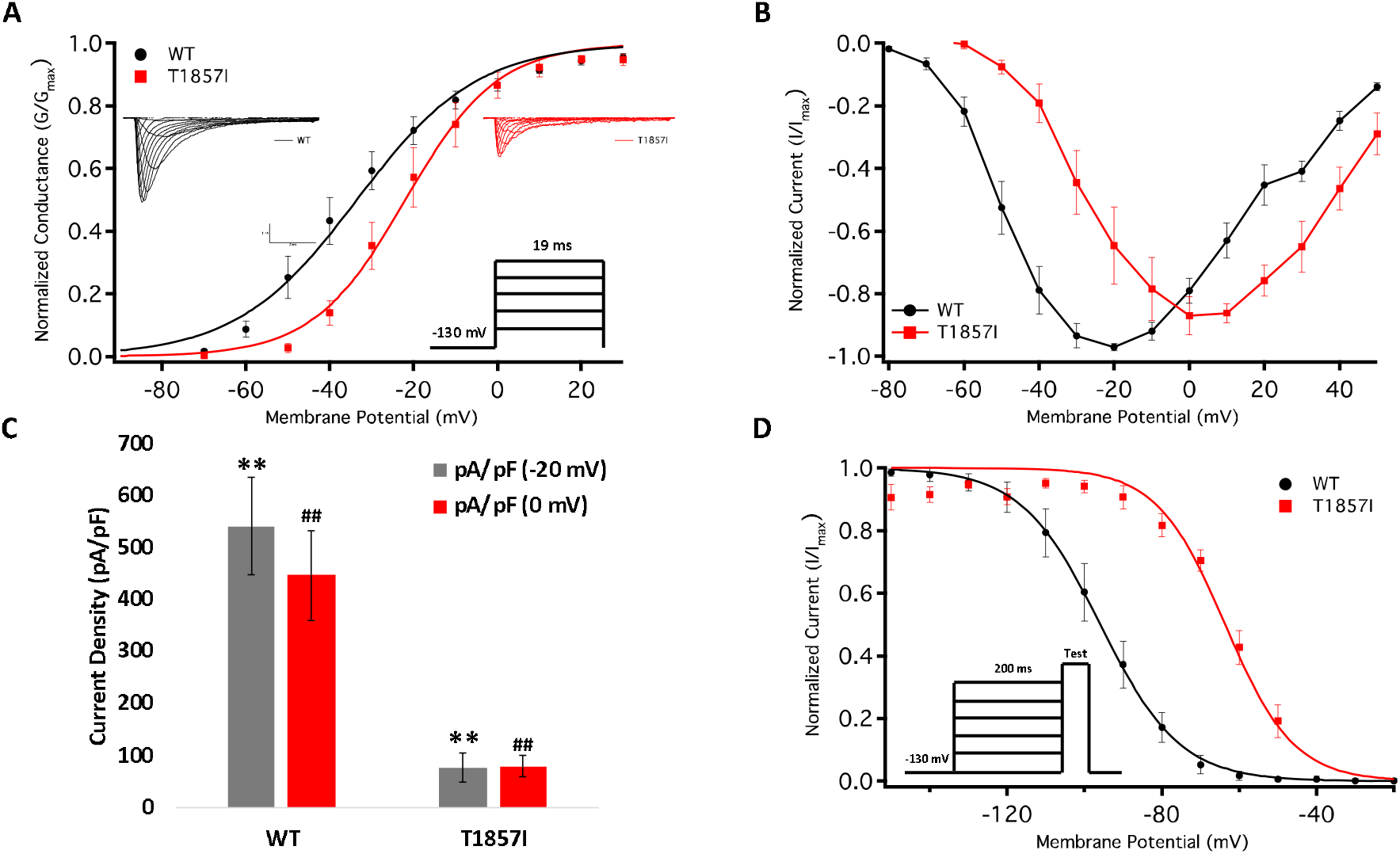
– Comparison of the Voltage-Dependence of Activation/Fast Inactivation between WT and T1857I. (**A**) Voltage-dependence of activation as normalized conductance plotted against membrane potential. Sample macroscopic sodium currents elicited by depolarizations between −100 and + 80 mV. (**B**) Normalized current and voltage relationship. (**C**) Average current (Y-axis) density of WT and T1857I. (**D**) Voltage-dependence of steady-state fast inactivation as normalized current plotted against membrane potential.

### T1857I Decreases Peak Current Density

We measured current density from the ratio of peak current amplitude to the cell membrane capacitance (pA/pF) (**Figure 2C**). As variant activation curve was right shifted compared to WT, we measured the peak current densities at −20 mV (where WT elicits maximal current) and 0 mV (where T1857I elicits maximal current (**Figure 2B**)). Our results suggested that the T1857I variant channels had significantly smaller current densities than WT at both potentials (p<0.0001) (**Figure 2C; Table S2**).

### T1857I Depolarizes Steady-State Fast Inactivation (SSFI)

We measured the voltage-dependence of SSFI using a standard 200 ms pre-pulse voltage protocol. Normalized current amplitudes were plotted as a function of pre-pulse voltage (**Figure 2D; Table S3**). Our results indicate T1857I significantly destabilizes the V_1/2_ (p<0.0001), but not the z (p>0.05) of the voltage-dependence of steady-state fast inactivation. This indicates that at any given potential the variant channels are less likely to fast inactivate, suggesting an increased excitability.

### T1857I Slows Recovery from Inactivation

One benchmark of Nav is the rate at which they recover from inactivation. To measure recovery, we held Nav1.5 at −130 mV to ensure channels were fully available, then pulsed the channels to 0 mV for 500 ms and allowed different time intervals at −130 mV to measure recovery as a function of time (**Figure 3A; Table S4**). Our results indicate that T1857I variants have a relatively slowed rate of recovery from fast inactivation (p<0.05).

**Figure 3.**
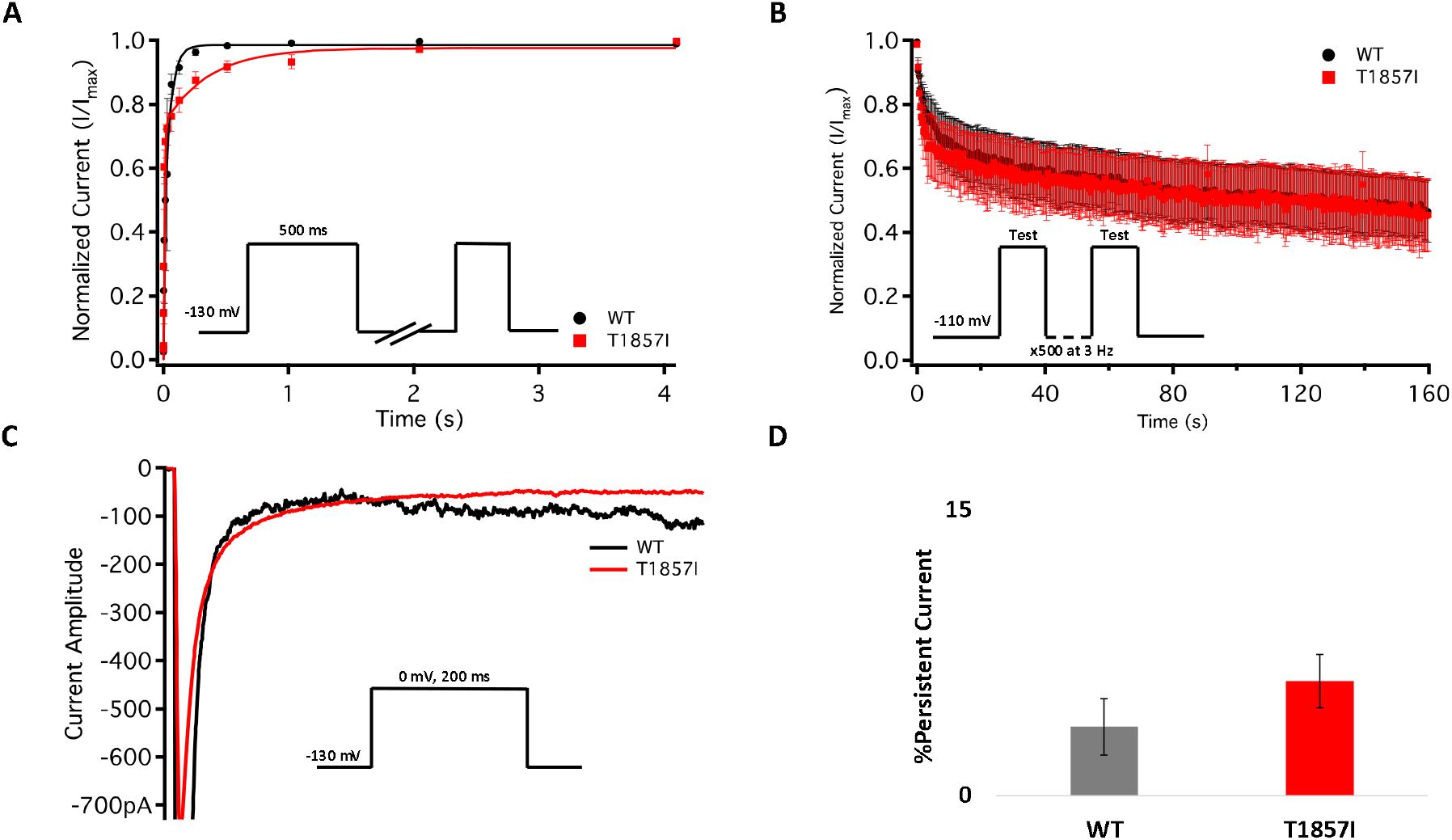
– Comparison of Fast Inactivation Recovery, Use-Dependence, and Persistent Currents. (**A**) Recovery from fast inactivation. (**B**) Use-dependent inactivation at 3Hz. Show normalized current decay plotted as a function of time. (**C**) Representative normalized current traces of late currents in WT and T1857I. (**D**) Average late sodium current as a percentage of peak sodium current.

### T1857I Does Not Alter Use-Dependent Inactivation

To understand the effects of elevated heart rate on channel function, we elicited use-dependent inactivation at 3 Hz. Normalized currents from 3 Hz use-dependent inactivation protocols are plotted against time in **Figure 3B** for both the channel variants. We found that there are no significant differences between WT and T1857I in use-dependent inactivation rates (p>0.05) (**Table S5**).

### T1857I Does Not Alter Late Currents, Increases Window Currents

Total sodium current can be divided into two components: peak (INaP) and late (INaL) currents. Whereas INaP refers to the maximum amount of sodium ions going through the channels during the open state, the smaller INaL is a manifestation of destabilized fast inactivation and has been shown to underlie LQT syndromes^11,19–21^. We show representative normalized current traces for both channels across all conditions (**Figure 3C; Table S6**). We measured the percentage of INaL by dividing the maximum INaL between 145 ms and 150 ms by INaP and found that there is no significant difference in the percentage of late currents between WT and T1857I. This is consistent with the normal QTc observed in the patient.

Changes in the voltage dependence of both steady-state activation and fast inactivation may lead to differences in the window current. The differences can be measured using a ramp protocol in which the voltage is maintained at a hyperpolarized potential and then ramped up to positive potentials (from −130 mV to 20 mV at 0.3 mV/ms)^19^. Consistent with the large right-shift of the SSFI curve, we found that T1857I has an increased sodium window current compared with WT-Nav1.5 (**Table 1**).

**Table 1-.** Window Current as A Percentage of Peak Current (n = 7-19)

| Channel Type | Mean % ± SE |
| --- | --- |
| WT | 1.7 ± 0.3<sup>a</sup> |
| T1857I | 7.8 ± 1.1 |

<sup>a</sup> taken from <sup>19</sup>

### T1857I Causes Afterdepolarizations in the O’Hara–Rudy Model of Ventricular Cardiac Excitability

We used the O’Hara–Rudy model to simulate cardiac action potentials (AP) and measure the effect of the T1857I mutation^15^. The control results from the patch clamp experiments were adjusted to the original model parameters and the subsequent magnitude shifts in the simulations of the mutation were performed relative to the original model parameters (**Figure 4**).

**Figure 4.**
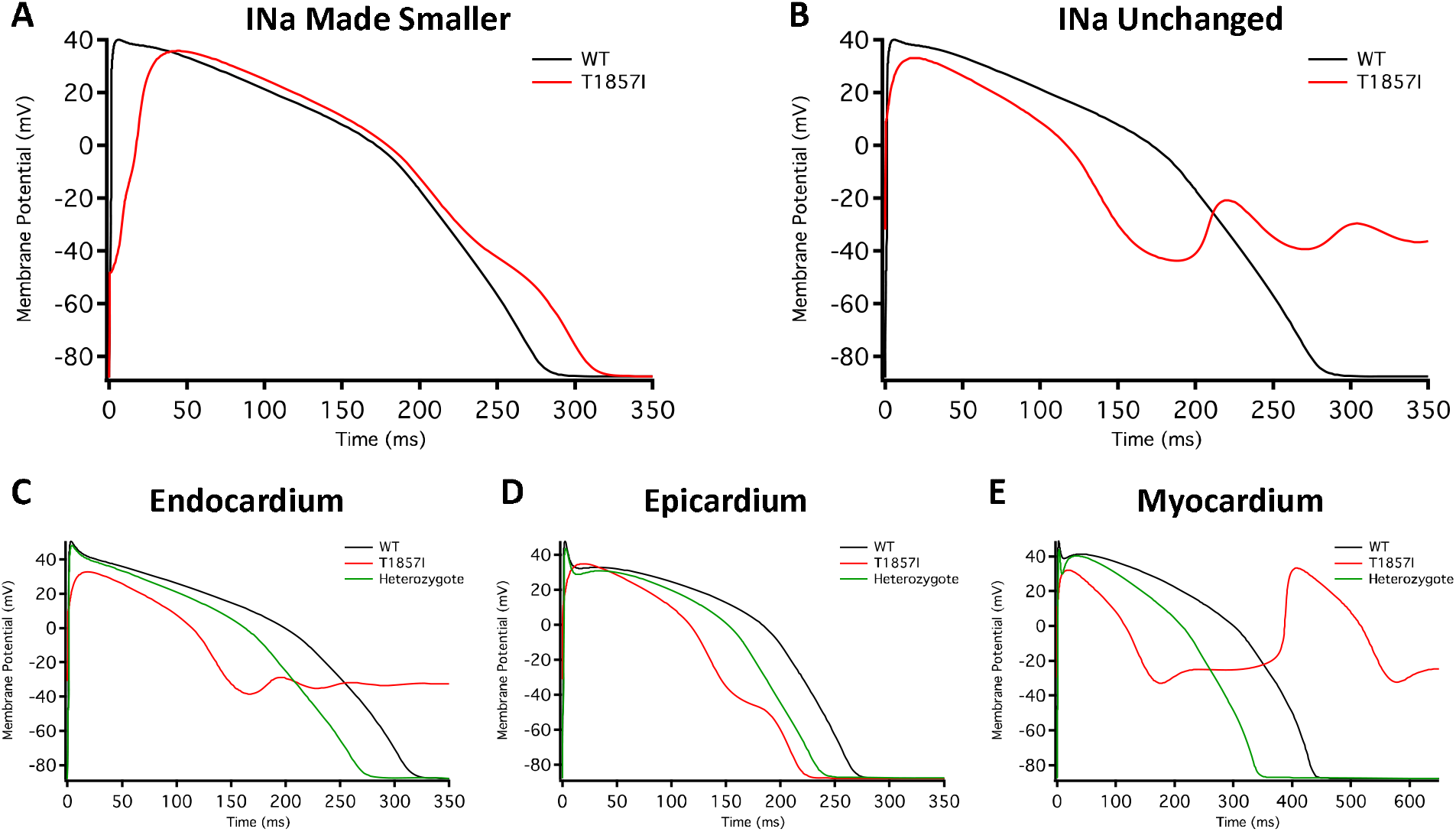
– O’Hara-Rudy-Based Ventricular Action Potential Model. Comparison of the action potential between WT and T1857I. The changes to conductances were based on experimental data obtained from patch-clamp results. (**A**) Changed gNa, (**B**) Unchanged gNa. (**C-E**) Endocardium/epicardium/myocardium simulations with WT, T1857I, and Heterozygote (unchanged gNa).

The smaller current densities in T1857I (**Figure 2C**) could have two possible underlying causes: 1) the variant biophysically alters and reduces the sodium conductance, 2) the variant construct has a smaller expression/trafficking, hence the smaller sodium conductance in patch-clamp experiments is an artificial effect. Therefore, we performed two sets of simulations for T1857I. In **Figure 4A**, we show an AP simulation where we reduced the sodium conductance by ~6 fold, which is consistent with the current density data shown in **Figure 2C**. The results from this condition show reduction in the upstroke velocity and peak amplitude of phase 0, and a slight afterdepolarization hump that starts at ~250 ms. In **Figure 4B**, we left the sodium conductance unchanged in T1857I. In this simulation, a more pronounced afterdepolarization was observed starting at ~225 ms. Overall, both sets of simulations suggest that T1857I causes afterdepolarizations, which could lead to cardiac arrhythmias.

To examine possible variant effects on different ventricular cell types, we compared simulated APs between endocardium, epicardium, and myocardium. Afterdepolarizations are observed in all three cell types (**Figure 4C-E**).Also, as our patients are heterozygotes (maternal inheritance of the variant allele), we performed all simulations with a heterozygous assumption (i.e. 50% of the channels have the variant phenotype and 50% are WT). The AP morphology of the heterozygous simulations appear less exaggerated than the pure T1857I conditions, and therefore may be a better representation of ventricular excitability in our patients, congruent with the normal QRS duration on baseline ECG in both patients.

### T1857I Causes Slight GOF in Atrial Grandi Model of Cardiac Excitability

To investigate possible T1857I effects on atrial excitability, we ran atrial AP simulations using the Grandi model^17^ (**Figure 5A-D**). Similar to the ventricular APs, reducing the sodium conductance lowered the AP peak and slightly prolonged the APD (**Figure 5A**). However, in the simulations in which the sodium conductance remained unchanged, the variant effects were minimal (**Figure 5B**). Furthermore, there were virtually no differences in the variant effect between atrial epicardium and endocardium simulations (**Figure 5C-D**), and these effects became even more subtle in heterozygote conditions. Overall, based on the Grandi model, the effect of T1857I on atrial excitability is predicted to be minimal.

**Figure 5.**
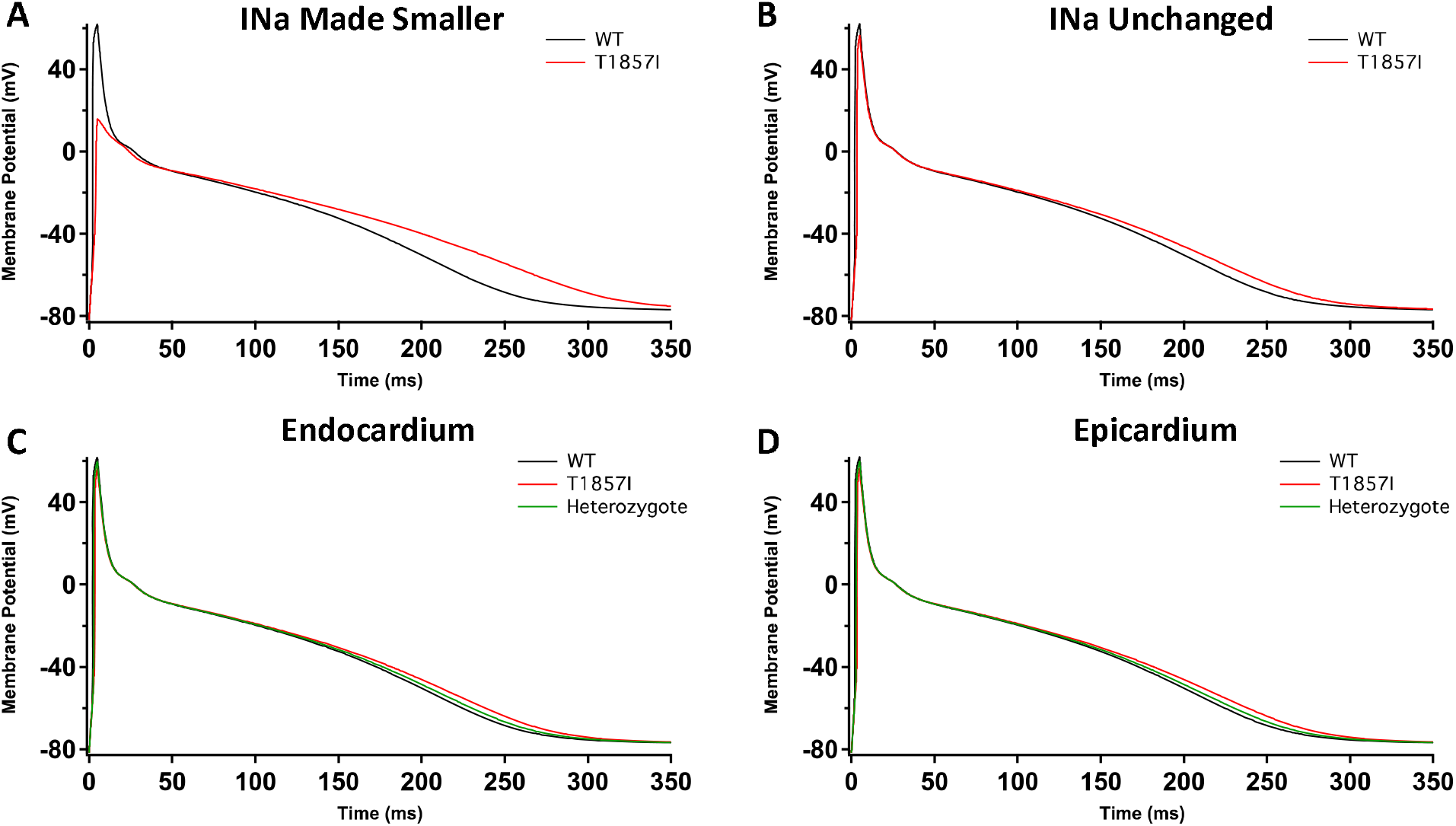
– Grandi-Based Atrial Action Potential Model. Comparison of the action potential between WT and T1857I. The changes to conductances were based on experimental data obtained from patch-clamp results. (**A**) Changed gNa, (**B**) Unchanged gNa. (**C-D**) Endocardium/epicardium simulations with WT, T1857I, and Heterozygote (unchanged gNa).

## Discussion

We identified a novel variant, T1857I, explored its effects on the biophysical properties of Nav1.5, and used the results to simulate cardiac action potentials. Our results suggest that the voltage-dependence of both activation and inactivation are right-shifted. The right-shift of activation suggests a loss-of-function, in contrast the larger right-shift of inactivation suggests a gain-of-function in this channel. We also found that the recovery from fast inactivation was slowed. To determine the effects of gating shifts in T1857I, we first introduced the relevant experimental data into the ventricular O’Hara-Rudy model, which suggests afterdepolarizations may result. We then performed atrial AP modeling using the Grandi model, which suggested minimal variant effects. Thus, we conclude that T1857I likely causes a net gain-of-function in the Nav1.5 gating, with an increased window current and resultant ventricular tissue afterdepolarization, which could lead to arrhythmias. These biophysical properties of afterdepolarization, unaltered late Na^+^ current and unchanged AP duration are respectively consistent with the clinical phenotype of the probands characterized by frequent and polymorphic ventricular ectopy and tachy-arrhythmia, normal QTc and QRS duration.

The inward sodium current through Nav1.5 is the principal determinant of V_max_ inside the myocardium. The functional differences in human atrial and ventricular sodium current biophysical properties were suggested to be minimal^22–25^. Although Nav1.5 is the predominant Nav channel in the heart, the transcript for the sodium channel auxiliary β1-subunit is more highly expressed in the atrium^26^. Recent structural studies determined the structural basis for a reduced modulation of Nav1.5 by β1-subunits. They suggested that the sodium current amplitude differences between atrial and ventricular cells are unlikely to be significantly affected by β1-subunits expression levels^11,13^. Additionally, reports in canine atrial cells suggest that the atria have a larger sodium current density and a hyperpolarized inactivation compared to ventricles^27^. Also, due to the depolarized resting membrane potential, there is less sodium channel availability in the atria compared to the ventricle, and hence, the resulting sodium current underlying the atrial action potential is smaller in atrial cells^28^. In contrast to the sodium current, several other ionic currents display more pronounced differences. These differences underly the difference in action potential morphology between these cell types^29^. These differences may also be the reason for the less pronounced T1857I experimental simulated effects in atria compared to ventricles (**Figure 4-5**).

The sodium/calcium exchanger (NCX) plays a vital role in maintaining cellular calcium levels. NCX takes advantage of higher extracellular sodium concentrations to pump calcium out of the cytosol. However, the NCX also functions in reverse mode^4,30^. The human atrial action potential is also modulated by intracellular calcium concentrations, which can directly affect calcium channel inactivation^31^ that modulates the timing and size of NCX current^32,33^. Our results suggest that the most notable biophysical property of T1857I is its ~30 mV right-shifted SSFI. This large shift increases the overlap (window current) between the SSFI and activation curves in T1857I. Indeed, direct measurements of this window current by a ramp protocol further highlights this increase (**Table 1**). The elevated window current may explain the afterdepolarizations observed in our AP simulations (provide figure). Given this SSFI curve, at any given membrane potential, more sodium influx into the cytosol is expected. In turn, this may induce reverse mode NCX activity leading to Ca^2+^ entry and give rise to afterdepolarizations in both atrial and ventricular myocytes. This hypothesis merits further investigations through measurement of NCX activity in cardiac myocytes.

The phenotypic expression of T1857I is similar to that of R222Q, which underlies multifocal ectopic Purkinje-related premature contractions^34^. Both disorders include atrial arrhythmias, premature ventricular contractions, and may result in SCD. However, the biophysical underpinnings of the two syndromes are quite different. Although both R222Q and T1857I cause a depolarizing shift in the voltage dependence of activation, our results with T1857I also show a depolarizing shift in steady-state fast inactivation, whereas R222Q causes a small hyperpolarizing shift in inactivation. Furthermore, the *in-silico* action potentials have different morphologies arising from the two mutations. These differences underscore the novelty of our clinical and biophysical observations related to T1857I.

## Conclusions

In conclusion, we report a novel *SCN5A* mutation encoding T1857I where afflicted probands present with clinically characterizable incidence of atrial and ventricular arrhythmias and multiple familial sudden deaths. The biophysical characterization of this variant indicates a mixture of gain- and loss-of-function effects. Computer simulations using the experimental data suggest afterdepolarizations as the net result. These mechanistic findings are consistent with the clinically observed cardiac arrhythmias in the affected probands.

## Data Availability

All data are freely available.

## Conflict of Interest

None. The authors declare that this research was conducted in the absence of competing interests.

## Authors Contributions

M-RG performed all functional experiments, AP modeling, data analysis, figure making, wrote manuscript. PCR conceived the experiments and revised the manuscript critically. JA and CAE performed all clinical work. All co-authors edited the manuscript.

## Funding

This work was supported by grants from Natural Science and Engineering Research Council (NSERC) of Canada and the Rare Disease Foundation to PCR and M-RG (CGS-D: 535333-2019 & MSFSS: 546467-2019), a MITACS Accelerate fellowship in partnership with Xenon Pharma, Inc to M-RG (IT10714).

## Supporting Information

**Table S1-.** Conductance (n = 6-9)

| Channel Type | Mean $V_{1/2} \pm SE$ (mV) | Mean $z \pm SE$ (slope) |
| --- | --- | --- |
| WT | $-35.8 \pm 3.8$ | $2.3 \pm 0.3$ |
| T1857I | $-20.2 \pm 3.1^*$ | $3.1 \pm 0.2^*$ |

\* Statistical significance (p-value provided in text)

**Table S2-.** Current Density (n = 7-11)

| Channel Type | Mean density (-20 mV) $\pm SE$<br>(pA/pF) | Mean density (0 mV) $\pm SE$<br>(pA/pF) |
| --- | --- | --- |
| WT | $543 \pm 64$ | $447 \pm 57$ |
| T1857I | $77 \pm 51^*$ | $80 \pm 45^*$ |

\* Statistical significance (p-value provided in text)

**Table S3-.** SSFI (n = 6-9)

| Channel Type | Mean $V_{1/2} \pm SE$ (mV) | Mean $z \pm SE$ (slope) |
| --- | --- | --- |
| WT | $-96.1 \pm 3.0$ | $-2.8 \pm 0.3$ |
| T1857I | $-61.6 \pm 2.4^*$ | $-2.8 \pm 0.2$ |

\* Statistical significance (p-value provided in text)

**Table S4-.**
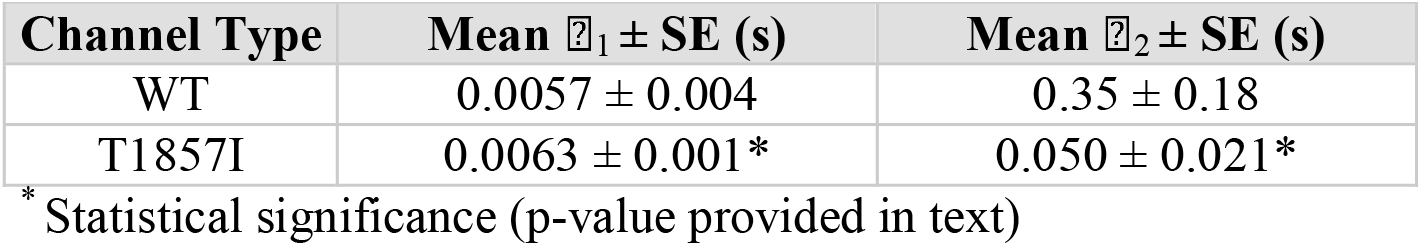
Fast Inactivation Recovery (n = 3-6)

**Table S5 -.** 3Hz (n = 7-8)

| Channel Type | Mean $\tau \pm SE$ (s) |
| --- | --- |
| WT | $12.3 \pm 3.0$ |
| T1857I | $9.0 \pm 2.8$ |

**Table S6.** –INaL (n = 3-7)

| Channel Type | Mean % $\pm SE$ (mV) |
| --- | --- |
| WT | $3.6 \pm 1.5$ |
| T1857I | $6.0 \pm 1.0$ |

## Notes

### Competing Interest Statement

The authors have declared no competing interest.

### Author Declarations

University of Alberta IRB ethics approval.

